# Colorectal cancer screening based on predicted risk: a pilot randomized controlled trial

**DOI:** 10.1101/2024.03.15.24304344

**Authors:** Ekaterina Plys, Jean-Luc Bulliard, Aziz Chaouch, Marie-Anne Durand, Luuk A. van Duuren, Karen Braendle, Reto Auer, Florian Froehlich, Iris Lansdorp Vogelaar, Douglas A. Corley, Kevin Selby

## Abstract

**Background & Aims:** Colorectal cancer (CRC) screening relies primarily on colonoscopy and fecal immunochemical testing (FIT). Aligning utilization of these options with individual CRC risk (i.e. personalized screening) may maximize benefit with lower risks, individual burdens, and societal costs. We studied the effect of communicating personalized CRC risk and corresponding screening recommendations on appropriate screening uptake in an organized screening setting.

**Methods:** Pilot randomized controlled trial among residents aged 50-69 years old not yet invited for screening in Vaud, Switzerland. The intervention was a mailed brochure communicating individual 15-year CRC risk and corresponding screening recommendation. The control group received a brochure comparing FIT and colonoscopy. The primary outcome was self-reported risk-appropriate screening (FIT if <3% risk, FIT or colonoscopy if ≥3% and <6%, colonoscopy if ≥6%), assessed by a mailed questionnaire at 6 months. A secondary outcome was overall screening uptake.

**Results:** Of 5396 invitations, 1059 people responded (19%), of whom 258 were randomized to intervention and 257 to control materials (average 15-year risk 1.4% (SD 0.5), age 52.2 years (SD 2.2), 51% women). Risk-appropriate screening completion was 37% in the intervention group and 23% in the control group (absolute difference 14%, 95%CI 6%-22%, p<0.001). Overall screening uptake was 50% in the intervention and 49% in the control group (absolute difference 1%, 95CI −7%−10%, p=0.758).

**Conclusions:** In a population not known to be at elevated CRC risk, brochures providing personalized CRC risk and screening recommendations improved risk-appropriate screening without impacting overall screening uptake. This approach could be helpful for aligning screening methods, risks, and benefits with cancer risk. **Trial registration:** Clinicaltrials.gov NCT05357508.

**What You Need to Know:** *Background:* Colorectal cancer can be effectively prevented by screening using colonoscopy or fecal immunochemical test (FIT). Optimizing use of colonoscopy resources is crucial to reduce screening burden for patients and society.

*Findings:* After reading our intervention brochure, participants were 14% more likely to choose the screening test appropriate to their risk level. This result did not impact overall screening participation.

*Implications for patient care:* Risk-based screening recommendations for FIT or colonoscopy could be a means of better allocating colonoscopy resources in countries relying heavily on colonoscopy for screening, thus decreasing the burden of CRC screening for low-risk participants.

## Introduction

Colorectal cancer (CRC) is the third most common cancer in men and second most common in women, causing approximately 700 000 deaths worldwide every year [1]. The long pre-clinical development of the disease allows for screening to reduce CRC incidence and mortality [2, 3]. In Switzerland 48% of the population aged above 50 was up to date with screening in 2017, and 43% had had a screening or diagnostic colonoscopy within the last 10 years [4]. Although colonoscopy is preferred because of its high sensitivity for CRC and advanced adenomas [5], there is no direct evidence for its superiority compared to the Fecal immunochemical test (FIT) in an organized screening setting to detect cancers [6] and prevent CRC mortality [7], especially among individuals at low or average risk of cancer. Moreover, colonoscopy exposes individuals to potential complications [2]. When overused as a screening method, it occupies gastroenterologist resources, resulting in longer waiting time for the individuals with symptoms and higher health-related costs for society [8].

Personalized CRC screening could decrease overuse of colonoscopy by reserving this method for individuals at high risk and orienting others to FIT. Personalized screening includes estimating individual risk for CRC and providing screening recommendations appropriate to that risk level. This could diminish potential harm related to screening, namely the burden of colonoscopy preparation, risk of side-effects, and inappropriate expenditure [9]. However, it could also cause anxiety and a sense of inevitability when participants are communicated their personal risk by mail, without a healthcare provider.

The impact of personalized screening recommendations on appropriate screening is not clearly understood. Most studies have tested its influence on overall screening uptake, with multiple studies and a meta-analysis concluding that personalized risk information results in little to no effect [10–14]. Only two studies focused on appropriate screening, meaning use of colonoscopy by those at high risk and non-invasive tests by those at low risk [15, 16]. It appears that communicating risk alone without personalized screening recommendations is insufficient to influence individuals’ decision about screening test [16]. Emery et al [15] showed an increase of 21% in risk-appropriate screening after communicating risk-based recommendations. However, risk and screening recommendations were communicated to participants by health providers in primary care, which is a challenging strategy in an organized screening setting.

The primary aim of this trial was to study whether communicating individual CRC risk and screening recommendations with written materials has an effect on appropriate screening uptake at six months. We hypothesized that participants at low risk would be more likely to undergo a FIT and participants at high risk a colonoscopy.

## Methods

This study followed the CONSORT Guidelines for reporting outcomes in trial reports [17]. The trial was registered prior to inclusion of the first participant and later published [18].

### Trial design

This was a monocentric, two-arm randomized controlled superiority trial with participants randomized 1:1 in the intervention and the control arms. The trial was nested in the CRC screening program of the canton of Vaud, Switzerland.

### Participants

Recruitment took place between June and September 2022 in the canton of Vaud. We included people aged between 50 and 69 years old who had not yet been invited to the cantonal organized CRC screening program. Individuals were excluded if they had symptoms suggestive of CRC, personal history of CRC, advanced adenoma or inflammatory bowel disease, genetic syndromes representing high risk for CRC (i.e., Lynch syndrome), if they were up-to-date with screening (colonoscopy within 9 years or FIT within 1.5 years) or expected to leave Switzerland during the 6-months follow-up. These exclusion criteria were verified using our self-administered recruitment questionnaire. See Figure 1 and Supplementary Figure 1 for details.

**Figure 1.**
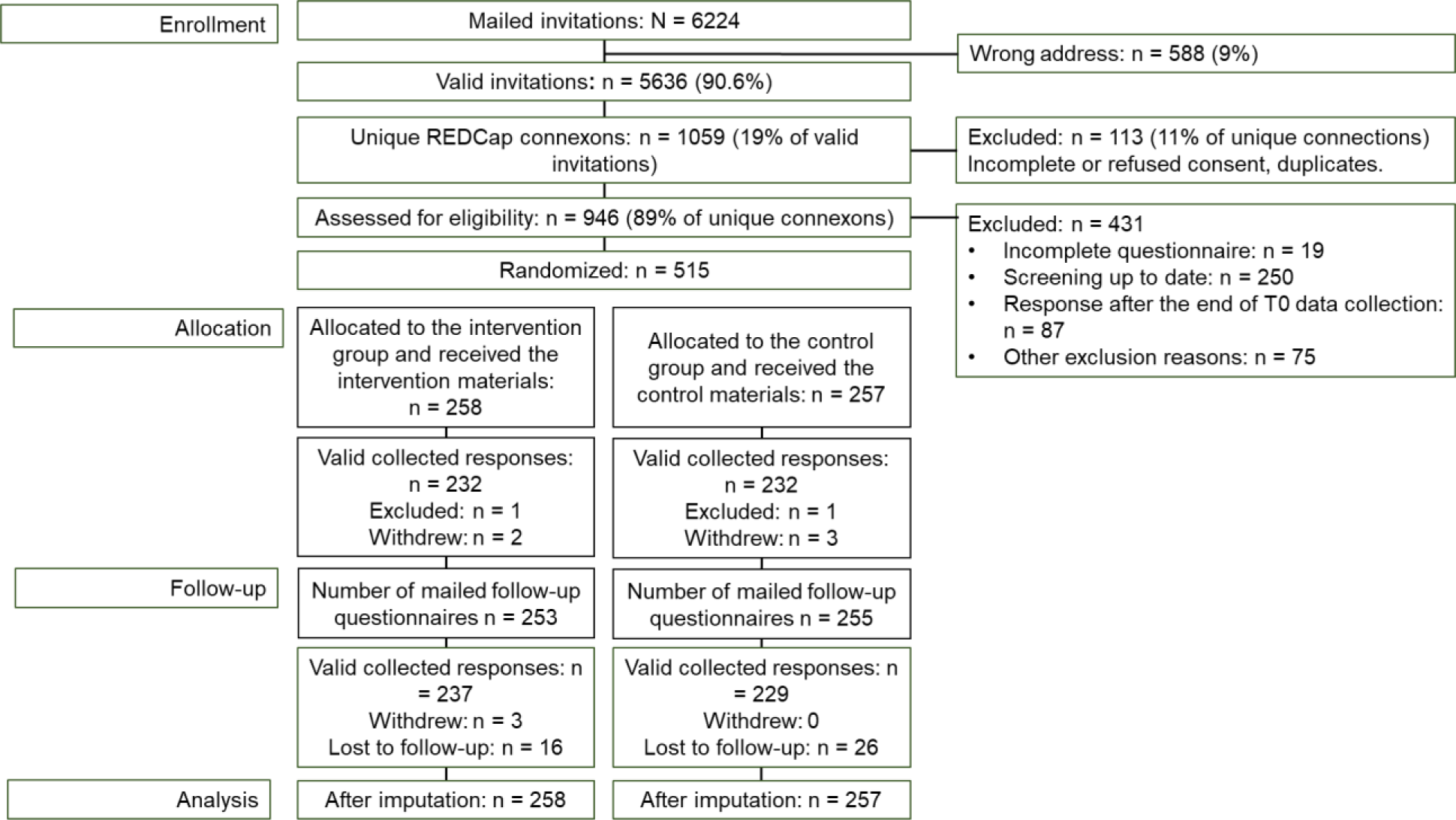
Flowchart of participants

### Intervention and comparator

The intervention consisted of mailing participants a brochure containing their 15-year CRC risk and corresponding screening recommendations. The individuals at low risk (<3%) were recommended the FIT, those at high risk (≥6%) were recommended colonoscopy, and those at moderate risk (3-6%) were offered a choice between the FIT and colonoscopy.

The intervention was based on the health belief model [19]. This model posits that people are more likely to adopt a health-protecting behavior if they believe they are susceptible, the illness is severe, the recommended behavior is efficient, and when they feel higher self-efficacy [19]. It was expected that, after the intervention, people at high risk would feel more susceptible and would therefore complete colonoscopy, whereas people at low risk would feel less susceptible and prefer the non-invasive FIT. Screening recommendations were expected to increase self-efficacy. Brochures for those at low risk informed them that zero risk didn’t exist and strongly recommended a screening test. As there are no strong recommendations for the individuals at moderate risk, the brochure presented FIT and colonoscopy as equally reasonable. The control brochure was the standard brochure used by the Vaud screening program, which presented the advantages and disadvantages of both FIT and colonoscopy. It did not include personalized risk or risk-based screening recommendations. For more information, see Supplemental materials and our study protocol [18].

An individual’s personal 15-year CRC risk score was calculated using the QCancer-colorectal risk calculator [20]. This open-source tool developed and validated in the United Kingdom assigns individuals aged 40 to 69 years old to the correct risk group 66-70% of the time [21, 22]. For this study, the QCancer algorithm was modified slightly by excluding postal code, ethnicity, ulcerative colitis, and colonic polyps as risk factors (see [18] for details). Based on the obtained risk score, individuals were divided in three risk levels: low risk (<3%), moderate risk (3% - 6%), and high risk (≥6%), based in part on the 3% threshold of the BMJ Rapid Recommendation [23].

### Recruitment and data collection procedures

The study included three phases. At T0, 6200 invitations with consent and the recruitment questionnaires were mailed, which allowed us to verify eligibility for the study and calculate CRC risk. At T1, right after receiving consent, the participants received intervention or control materials, questionnaire 2 and an information sheet to facilitate discussion about screening with their general practitioner or pharmacist. At T2, 6 months after the intervention, screening behavior was measured using the follow-up questionnaire. Because mailings and return can take several weeks, responses were accepted up to 8 months after the intervention. The consent and all three questionnaires were available in electronic and paper form on the REDCap platform [24]. All study documents were written in French. For more details on the procedure, see our study protocol [18].

### Outcome measures

Our primary outcome was self-reported appropriate screening uptake measured 6 months after the intervention. Screening was considered appropriate when participants at low risk completed FIT, those at high risk completed colonoscopy, and those at moderate risk completed colonoscopy or FIT. Since waiting time for colonoscopy can be long [8], a colonoscopy appointment was considered as a completed test.

### Secondary outcomes

Overall screening participation (at T2) was calculated as the proportion of individuals who completed any CRC screening test. Anxiety related to the printed materials (at T1) was assessed using six items adapted from the Spilberger’s State-Trait Anxiety Inventory [25]. Participants were asked to respond to these items after having read the brochure. See Supplementary materials for the questionnaires in French and English.

### Quality assurance, citizen engagement, and ethical considerations

Only validated questionnaires or questionnaires pre-tested in previous studies were used. After entering data in REDCap [24], 20% of responses collected using paper questionnaires were double checked. Five individuals of the target population (a citizen advisory group) were involved in development of the materials for participants, and in interpretation and dissemination of the results. The trial was monitored by an institutional monitoring team and received approval from the Ethics committee of the canton of Vaud on March 2, 2022 (project ID 2021-02431).

### Sample size

Estimations of the sample size were explained in detail in our study protocol [18]. It was expected that 60% of the individuals eligible for screening would be at low risk, 10% at high risk, and 30% at moderate risk. Our intervention was expected to increase appropriate screening in low and high-risk participants, but not those at moderate risk. To detect a difference of 10% between groups with power of 80% and an alpha of 5%, we needed 393 individuals in each group. After considering attrition of about 10%, the final sample size was estimated at 440 individuals in each group (880 in total).

During recruitment, we realized that nearly all participants eligible for the study were at low risk, leading us to recalculate our sample size. With 95% of participants at low risk, we would need 451 participants in total to have 80% power to detect a 15% change in appropriate screening.

### Randomization

The REDCap automatic randomization module was programmed with the block factor varying between 4 and 8. Randomization was carried out after entering the risk score and risk level by clicking on the button “Randomize”. The trial team member who carried out the randomization was not blinded. Participants were told that the study aim was to compare two brochures on CRC screening. They, therefore, were not aware of group assignment. The trial statistician who conducted the primary outcome analyses was blinded to participants’ group assignments.

### Statistical methods

Intention to treat analyses were conducted. Randomization quality was tested using a 2-tailed chi-squared or Fisher exact tests for categorical variables; Student’s t-test or Manny-Whitney U tests were used for continuous variables. The primary outcome was analyzed using 2-tailed chi-squared test. Participants who refused screening and those who did not respond to the follow-up questionnaire were treated as not having done a screening test in both groups. Pre-specified subgroup analyses for the main outcome were performed by sex, nationality, education, occupation, French proficiency level, household size, family history of CRC and polyps using a proportion test. The overall participation was tested using 2-tailed chi-squared test. The other secondary analyses were conducted using 2-tailed t-test. Data analyses were performed using the Stata 16 [33] and R software packages [32].

### Important changes to methods after trial commencement

After trial commencement, several modifications to the protocol were made. Given the lower than expected response rate after the first wave of invitations in May – June 2022, 2000 more invitations were sent in August – September 2022. An additional letter was sent to the participants to communicate their identification code in the Vaud screening program as this information is mandatory to activate the participants’ personal file in the screening program database. The primary outcome’s data collection procedure was also slightly changed. In addition to the mailed questionnaire, the participants who had not responded were contacted by the phone, which allowed us to collect an additional 27 responses.

### Data Sharing

Deidentified individual participant data are available from the corresponding author on reasonable request.

## Results

### Participant characteristics

Of 6,200 mailed invitations, 5,396 went to a valid address, 1,059 individuals responded to the invitation letter (19.6% of valid addresses), and 946 signed the main consent form (17.5%). Among them, 318 were excluded because they did not meet inclusion criteria (Table 1), 87 because they responded after the end of the recruitment, 19 because they did not complete questionnaire 1, and seven because of an error at randomization (see Figure 1 and Table 1). The remaining 515 participants were randomized to the intervention (n=258) or the control group (n=257). The mean age was 52.2 years (SD=2.2) and the mean 15-year CRC risk was 1.4% (SD=0.5). Stratification by risk level showed that 98.1% (n=505) of the participants were at low risk, 1.9% (n=10) were at moderate risk and 0 at high risk. Sociodemographic characteristics, family history of polyps or CRC, and the baseline intention to be screened were similar between intervention and control groups (Table 2). Post-randomization, five withdrawals were registered in the intervention group and 3 in the control group. Seven participants withdrew without giving any reason and one because of insufficient knowledge of French. An additional two participants were excluded post-randomization at T1 (1 from the intervention and 1 from the control group) as they had done screening before receiving our printed materials. Finally, 42 participants (16 in the intervention and 26 in the control group) did not complete the follow-up questionnaire after reminders (see Figure 1). Participants without data for the primary outcome were assumed to not have completed a screening.

**Table 1.** Exclusion reasons.

| Exclusion reason | n | % |
| --- | --- | --- |
| Total excluded | 505 | 100 |
| Colonoscopy up-to-date | 223 | 44.2 |
| FIT up-to-date | 36 | 7.1 |
| FIT and colonoscopy up-to-date | 8 | 1.6 |
| Genetic risk | 42 | 8.3 |
| Inflammatory disease of the colon | 6 | 1.2 |
| Regular control for polyps | 58 | 11.5 |
| Serious disease that could prevent from participation in the study | 2 | 0.4 |
| Unexplained weight loss | 7 | 1.4 |
| Blood in stool | 32 | 6.3 |
| Unusual digestive disorders | 79 | 15.6 |
| Planning to leave Switzerland | 1 | 0.2 |

Note: multiple reasons for exclusion are possible.

**Table 2.** Participants baseline characteristics. FIT: fecal immunochemical test.

|  | Intervention group (n=258) | Control group (n=257) | Total |
| --- | --- | --- | --- |
| Sex |  |  |  |
| Women | 127/258 (49.2%) | 136/257 (52.9%) | 263/515 (51.1%) |
| Men | 131/258 (50.8%) | 121/257 (40.1%) | 252/515 (48.9%) |
| Age |  |  |  |
| n | 258 | 257 | 515 |
| Mean (SD), years | 52.1 (2.1) | 52.2 (2.4) | 52.18 (2.2) |
| Min/Max, years | 50-67 | 50-66 | 50-67 |
| Risk score |  |  |  |
| n | 258 | 257 | 515 |
| Mean (SD) | 1.3 (0.4) | 1.4 (0.6) | 1.38 (0.5) |
| Min/Max | 0.9-4.0 | 0.9-5.6 | 0.9-5.6 |
| Risk level |  |  |  |
| Low | 254/258 (98.4) | 251/257 (97.7) | 505/515 (98.1%) |
| Moderate | 4/258 (1.6) | 6/257 (2.3) | 10/515 (1.9%) |
| High | 0 (0.0) | 0 (0.0) |  |
| Nationality |  |  |  |
| Swiss | 199/258 (77.1) | 186/257 (72.4) | 385/515 (74.8%) |
| Other | 59/258 (22.9) | 71/257 (27.6) | 130/515 (25.4%) |
| Education |  |  |  |
| Compulsory education or less | 24/256 (9.4) | 16/254 (6.3) | 40/510 (7.8%) |
| Apprenticeship/Maturity | 94/256 (36.7) | 81/254 (31.9) | 175/510 (34.3%) |
| University/School of applied sciences | 135/256 (52.7) | 152/254 (59.8) | 287/510 (56.3%) |
| No response | 3/256 (1.2) | 5/254 (2.0) | 8/510 (1.6%) |
| Occupation |  |  |  |
| Employed | 227/256 (88.7) | 226/255 (88.6) | 453/511 (88.7%) |
| Unemployed | 25/256 (10.0) | 23/255 (9.0) | 48/511 (9.4%) |
| Retired | 3/256 (1.2) | 5/255 (2.0) | 8/511 (1.6%) |
| No answer | 1/256 (0.4) | 1/255 (0.4) | 2/511 (0.4%) |
| French level |  |  |  |
| Very good | 203/258 (78.7) | 203/255 (79.6) | 406/513 (79.1%) |
| Good or poor | 55/258 (21.3) | 52/255 (20.4) | 107/513 (20.9%) |
| Health literacy |  |  |  |
| High | 238/258 (92.2) | 238/257 (92.6) | 476/515 (92.4%) |
| Moderate or poor | 20/258 (7.8) | 19/257 (7.4) | 39/515 (7.6%) |
| Household size |  |  |  |
| Living with a partner or in family | 218/257 (84.8) | 216/257 (84.0) | 434/514 (84.4%) |
| Living alone | 39/257 (15.2) | 41/257 (16.0) | 80/514 (15.6%) |
| CRC family history |  |  |  |
| No | 251/258 (97.3) | 246/256 (96.1) | 497/514 (96.7%) |
| Yes | 7/258 (2.7) | 10/256 (3.9) | 17/514 (3.3%) |
| Polyp family history |  |  |  |
| No | 225/258 (87.2) | 230/254 (90.6) | 455/512 (88.9%) |
| Yes | 33/258 (12.8) | 24/254 (9.4) | 57/512 (11.1%) |
| Intention to be screened (1 = not at all, 5 = definitely) |  |  |  |
| Mean (SD) | 4.08 (1.2) | 4.02 (1.2) | 4.05 (1.2) |

FIT: fecal immunochemical test

### Primary outcome

At 6 months, 37% of the participants who received the intervention underwent risk-appropriate screening compared to 23% in the control group (14% absolute increase, 95% CI 6% to 22%). Results are represented in Table 3. Subgroup analysis by sex, education, French level, household size, and family history of CRC and polyps for the main outcome did not reveal statistically significant heterogeneity in the effect of the intervention between groups (see Supplementary table 2). There was a trend towards a greater effect of the intervention on women than men.

**Table 3.** Proportions of screening tests completed by participants at low and moderate risk by group.

|  | Low risk for CRC* |  | Moderate risk for CRC** |  | Overall |  |
| --- | --- | --- | --- | --- | --- | --- |
|  | Intervention group<br>n (%) | Control group<br>n (%) | Intervention group<br>n (%) | Control group<br>n (%) | Intervention group<br>n (%) | Control group<br>n (%) |
| Screening tests completed |  |  |  |  |  |  |
| Fecal Immunochemical Test (FIT) | 93 (36) | 57 (23) | 1 (25) | 1 (17) | 94 (36) | 58 (23) |
| Colonoscopy | 35 (14) | 67 (27) | 1 (25) | 1 (17) | 36 (14) | 68 (26) |
| No screening | 126 (50) | 127 (50) | 2 (50) | 4 (66) | 130 (50) | 131 (51) |
| Appropriate screening<sup>#</sup> | 93 (37) | 57 (23) | 2 (50) | 2 (33) | 95 (37) | 59 (23) |
| Non-appropriate screening | 161 (63) | 194 (77) | 2 (50) | 4 (67) | 163 (63) | 198 (77) |
| Overall participation in screening |  |  |  |  |  |  |
| Screened | 128 (50) | 124 (49) | 2 (50) | 2 (33) | 130 (50) | 126 (49) |
| Not screened | 126 (50) | 127 (51) | 2 (50) | 4 (67) | 128 (50) | 131 (51) |

\*Low risk defined as having a 15-year risk of CRC <3%
\*\*Moderate risk defined as having a 15-year risk of CRC ≥3% and <6%
<sup>#</sup>Appropriate screening defined as completing a FIT if at low risk for CRC and either a FIT or colonoscopy if at moderate risk

### Secondary outcomes

Overall screening uptake was 50% participation in the intervention and 49% in the control group (1% absolute difference, 95% CI -0.07, 0.1). Subgroup analyses by sex, education, French level, household size, and family history of CRC and polyps did not reveal significant differences between groups (see Supplementary table 3). Anxiety related to the intervention was low in both groups with the mean of 1.5 (SD=0.5) in the intervention group and 1.6 (SD=0.5) in the control group. The 2-tailed t-test revealed a t-statistic of −1.449 (df=444) and p=0.15. As the mean anxiety was low, no subgroup analyses were performed.

## Discussion

This trial aimed at studying the effect of communicating personalized CRC risk and risk-appropriate screening recommendations on participants’ screening behavior. Participants who received the intervention were 14% more likely to undergo risk-appropriate screening compared to control group participants, who received materials comparing colonoscopy and FIT as equal options. Overall screening participation was not affected by the intervention. These results support the use of personalized screening recommendations to better allocate colonoscopy resources in areas offering direct screening colonoscopy.

Our main results are in line with Emery et al. [15] in which personalized screening increased risk appropriate screening by 21%. Their intervention was delivered in person and followed by a consultation with a general practitioner who could order a fecal blood test or book a colonoscopy appointment. In our study, the intervention was a mailed brochure. Although the brochure explained how to access screening, additional steps were required to get a FIT kit or to book a colonoscopy appointment.

We believe that for a study with a simple design without face-to-face interactions and medical staff involvement, the increase in 14% in risk-appropriate screening is a promising result. This result could be because we gave a clear recommendation about screening along with individual risk information.

We had a greater impact on appropriate screening than Skinner et al. [16], who did not show a significant impact of communicated risk and multiple screening. We suppose that recommending more than one screening option could hamper decision-making and reduce screening uptake. In our study, we recommended FIT to the people at low risk. Colonoscopy was presented as an alternative option if the participant did not agree with our recommendation. We intentionally used short and tailored messages to avoid overwhelming the participants with the information and facilitate decision making.

In our study, overall screening uptake did not differ between groups which is in line with findings of Skinner et al [16], Rawl et al [10], and Yen et al [11]. It seems that communicating risk and screening recommendations is insufficient to enhance overall screening uptake. Other interventions like mailed FIT and patient navigation are more effective for increasing CRC screening rates [26].

There has been considerable uncertainty about how to incorporate CRC risk stratification models into organized CRC screening, including concerns that participants may not accept recommendations for less sensitive tests [27, 28]. Personalized screening could be incorporated into organized screening programs as a means of reassuring participants that FIT is appropriate for them. In Switzerland, between 2007 and 2017, fecal blood tests rates decreased from 11% to 5.2% and screening colonoscopy rates increased from 8.2% to 24.9% [4]. Reorienting people at low risk to FIT would help to offer screening with better risk-benefit balance and use resources more efficiently.

Anxiety related to the brochures was low and did not differ between groups. This is in line with the studies conducted by Smith et al [12] and van Erkelens [29] and the meta-analysis by Edwards [13], which did not reveal an increase in anxiety related to risk-based interventions.

### Strengths and limitations

This is one of the first studies examining the impact of personalized CRC risk and screening recommendations on appropriate screening. By nesting the study in an organized program, we were able to recruit a representative portion of the target population and to test an intervention suitable for widespread adoption. Quality of the collected data were ensured by the institutional trial monitoring team and the study steering committee. A citizen advisory group co-designed the study materials.

Our study did have limitations. Only individuals who had not yet been invited to the Program were recruited, which resulted in the inclusion of a younger population with a mean age of 52 years old. Because of the strong influence of age on CRC risk, we therefore had no high-risk participants and could not study screening behavior in people at high risk. In further studies, more effort should be done to recruit such populations.

Our study materials were available only in French. Although the intervention and control brochures were written in plain language and approved by the citizen advisory group, people with limited health literacy could have had difficulties participating.

Finally, our recruitment procedure required signing a consent form and completing questionnaires prior to receiving the study materials. This may have created a selection bias with only motivated individuals included in the study. This would limit our ability to extrapolate our participation rates to routine screening.

## Conclusion

This trial demonstrated that an intervention brochure that communicates CRC risk and appropriate screening recommendations can increase risk-appropriate screening uptake among people at low risk without impacting overall participation rate. Future research should evaluate the impact of this approach on high-risk individuals and the impact of personalized screening on the detection of advanced neoplasia. The current results will be valuable for screening programs in Switzerland and other settings relying primarily on screening colonoscopy to optimize CRC screening.

## Abbreviations

FIT: fecal immunochemical test
CRC: Colorectal cancer

## Data Availability

The data generated and analyzed during this study are available from the corresponding author on reasonable request.

## Acknowledgements

We would like to thank the 6 citizen partners who contributed throughout the study (Janine Chabloz, Simone de Rougemont, Ernest Frischknecht, Cruces Gaulis, Marie-Jo Magnin, et Jean-Marc Zaugg). We would like to thank Sarah Hediger and Angèle Bettens for their help preparing mailings. Finally, we would like to thank the participants for their trust and responses.

## Data Transparency Statement

The data set generated and analyzed during this study are available from the corresponding author on reasonable request.

---

**Supplementary figure 1.**
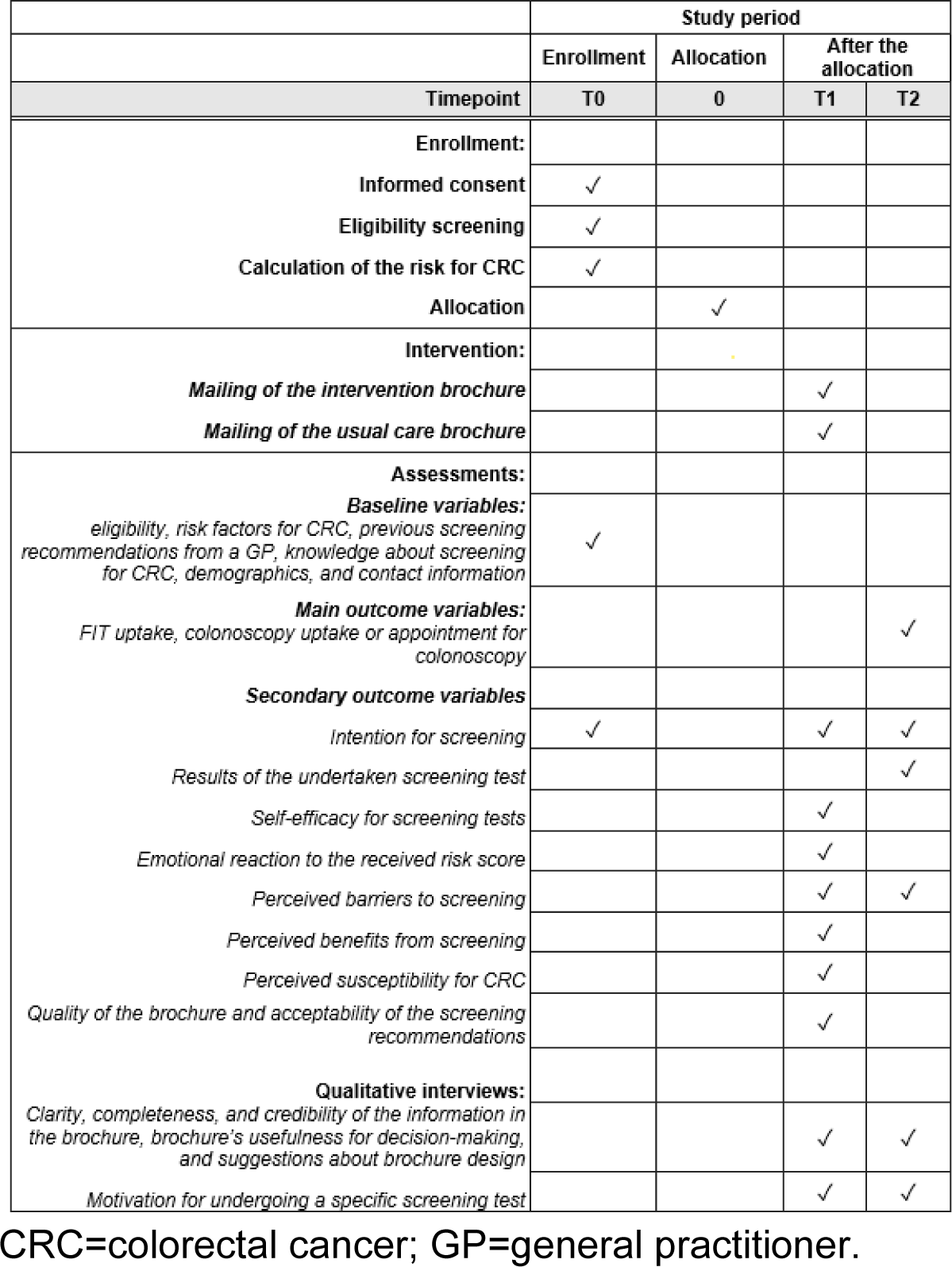
Schedule of enrolment, interventions, and assessments.

**Supplementary table 1.**
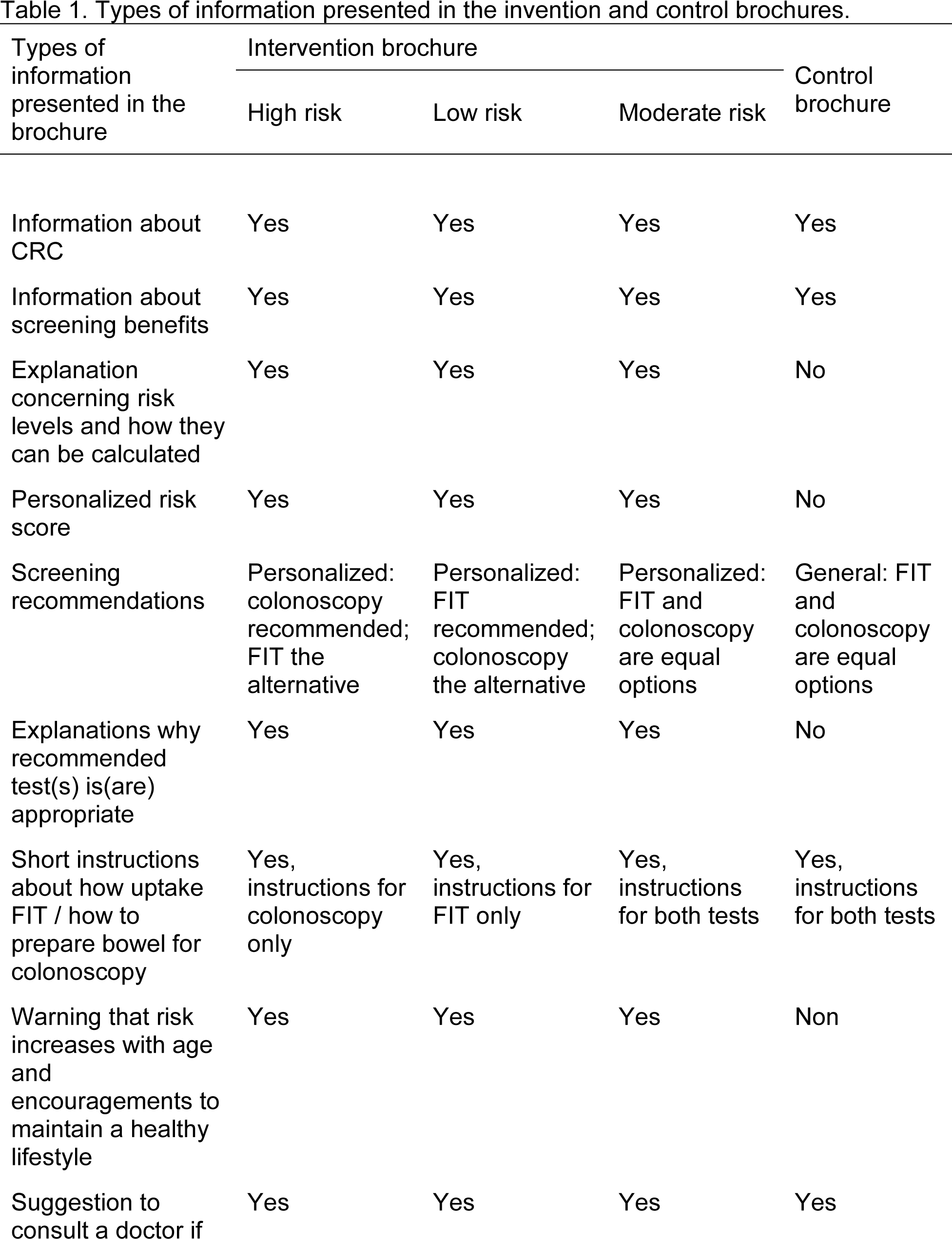
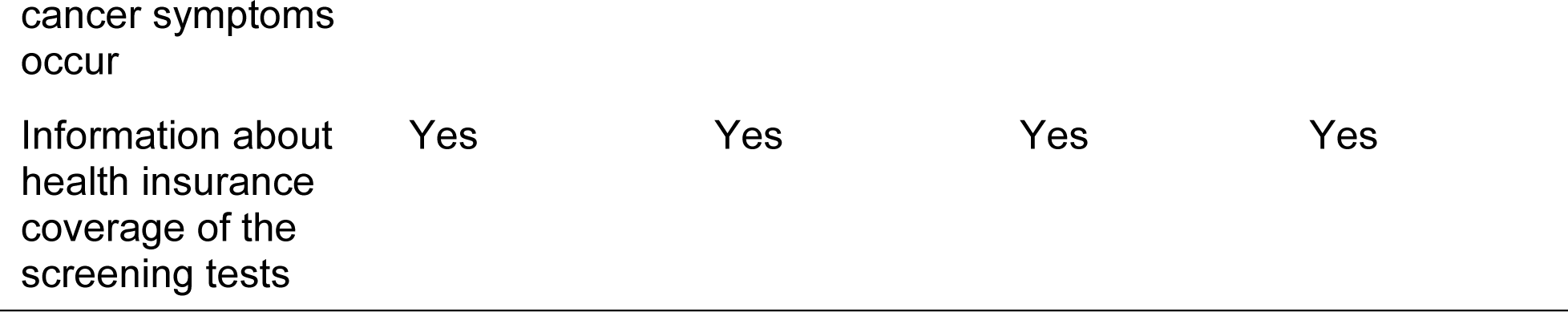
Types of information presented in the invention and control brochures.

**Supplementary table 2.** . Appropriate screening, subgroup analyses.

|  | Intervention group |  | Control group |  | Mean difference |  | p-value for interaction |
| --- | --- | --- | --- | --- | --- | --- | --- |
|  | Proportion | 95% CI | Proportion | 95% CI | Difference | 95% CI |  |
| Sex |  |  |  |  |  |  |  |
| Women (n=263) | 0.43 | 0.34, 0.51 | 0.22 | 0.15, 0.29 | 0.2 | 0.09, 0.32 | 0.13 |
| Men (n=252) | 0.31 | 0.23, 0.39 | 0.24 | 0.16, 0.32 | 0.07 | -0.04, 0.18 |  |
| Education |  |  |  |  |  |  |  |
| Compulsory school or less (n=40) | 0.33 | 0.14, 0.52 | 0.31 | 0.09, 0.54 | 0.02 | -0.27, 0.32 | 0.43 |
| Apprenticeship/Maturity/High school/University (n=462) | 0.37 | 0.31, 0.43 | 0.23 | 0.18, 0.29 | 0.14 | 0.06, 0.22 |  |
| French level |  |  |  |  |  |  |  |
| Very good (n=406) | 0.35 | 0.28, 0.42 | 0.22 | 0.16, 0.27 | 0.13 | 0.05, 0.22 | 0.71 |
| Good or poor (n=107) | 0.44 | 0.31, 0.57 | 0.25 | 0.13, 0.37 | 0.19 | 0.01, 0.36 |  |
| Household |  |  |  |  |  |  |  |
| Living with a partner or in family (n=434) | 0.38 | 0.31, 0.44 | 0.21 | 0.15, 0.26 | 0.17 | 0.08, 0.25 | 0.06 |
| Living alone (n=80) | 0.31 | 0.16, 0.45 | 0.34 | 0.2, 0.49 | -0.03 | -0.24, 0.17 |  |
| Family history of CRC and polyps |  |  |  |  |  |  |  |
| Yes (n=72) | 0.31 | 0.16, 0.45 | 0.18 | 0.05, 0.31 | 0.13 | -0.07, 0.32 | 0.98 |
| No (n=443) | 0.39 | 0.31, 0.44 | 0.24 | 0.18, 0.29 | 0.14 | 0.06, 0.23 |  |

**Supplementary table 3.** . Overall screening uptake, subgroup analyses.

|  | Intervention group |  | Control group |  | Mean difference |  | p-value for interaction |
| --- | --- | --- | --- | --- | --- | --- | --- |
|  | Proportion | 95% CI | Proportion | 95% CI | Difference | 95% CI |  |
| Sex |  |  |  |  |  |  |  |
| Women (n=263) | 0.56 | 0.42, 0.65 | 0.49 | 0.4, 0.57 | 0.07 | -0.05, 0.19 | 0.18 |
| Men (n=252) | 0.45 | 0.37, 0.54 | 0.5 | 0.41, 0.58 | -0.05 | -0.17, 0.08 |  |
| Education |  |  |  |  |  |  |  |
| Compulsory school or less (n=40) | 0.33 | 0.14, 0.52 | 0.63 | 0.39, 0.86 | -0.29 | -0.59, 0.01 | 0.07 |
| Apprenticeship/Maturity/High school/University (n=462) | 0.52 | 0.45, 0.58 | 0.5 | 0.43, 0.56 | 0.02 | -0.07, 0.11 |  |
| French level |  |  |  |  |  |  |  |
| Very good (n=406) | 0.51 | 0.44, 0.58 | 0.46 | 0.39, 0.53 | 0.04 | -0.05, 0.14 | 0.23 |
| Good or poor (n=107) | 0.49 | 0.36, 0.62 | 0.58 | 0.44, 0.71 | -0.08 | -0.27, 0.1 |  |
| Household |  |  |  |  |  |  |  |
| Living with a partner or in family (n=434) | 0.5 | 0.43, 0.56 | 0.47 | 0.4, 0.53 | 0.03 | -0.07, 0.12 | 0.41 |
| Living alone (n=80) | 0.54 | 0.38, 0.69 | 0.61 | 0.46, 0.76 | -0.07 | -0.29, 0.14 |  |
| Family history of CRC and polyps |  |  |  |  |  |  |  |
| Yes (n=72) | 0.59 | 0.44, 0.74 | 0.58 | 0.41, 0.74 | 0.01 | -0.22, 0.24 | 0.98 |
| No (n=443) | 0.49 | 0.42, 0.55 | 0.48 | 0.41, 0.54 | 0.01 | -0.08, 0.1 |  |

